# Alignment of contraceptive method attribute preferences and use among Kenyan women living with HIV

**DOI:** 10.1101/2025.08.21.25334189

**Authors:** Leah Hardenbergh, Aparna Seth, Barbra A. Richardson, Shiza Farid, Jenna I. Udren, Claire W. Rothschild, Nancy Mwongeli, Agnes K. Karume, Celestine Atieno, June Moraa, Jennnifer A. Unger, Daniel A. Enquobahrie, John Kinuthia, Alison L. Drake

**Affiliations:** Department of Epidemiology, University of Washington, Seattle, WA, USA; Department of Global Health, University of Washington, Seattle, WA, USA; Department of Biostatistics, University of Washington, Seattle, WA USA; FP2030, Washington, DC, USA; Department of Sexual and Reproductive Health, Population Services International, Washington, DC, USA; Department of Research and Programs, Kenyatta National Hospital, Nairobi, Kenya; Department of Obstetrics and Gynecology, Women and Infants Hospital, Warren Alpert Medical School of Brown University, Providence, Rhode Island, USA

**Keywords:** Contraception, contraceptive counseling, HIV, principal component analysis

## Abstract

**Objective:** Understanding alignment of contraceptive preferences and method selection among women living with HIV (WLWH) may improve contraceptive counseling. We examined whether method attribute preferences aligned with method attributes used among WLWH, and identified preference clusters.

**Study Design:** We used baseline data from WLWH enrolled in a cluster randomized controlled trial of a reproductive health counseling intervention in Kenya. Women using modern contraception at baseline were eligible (N=2,599). We used Poisson regression models to characterize 11 relationships between method attributes preferred vs used, and principal component analysis (PCA) to identify preferences clusters.

**Results:** Women who preferred methods that are long-acting (adjusted Prevalence Ratio [aPR]:1.63, 95% Confidence Interval [CI]:1.19-2.24), avoid daily dosing (aPR:1.11, 95% CI:1.07-1.16), permit self-discontinuation (aPR:1.32, 95% CI:1.14-1.52), and are concealable (aPR:1.06, 95% CI:1.01-1.12) were significantly more likely to use aligned methods. Attribute preferences clustered on three dimensions: (1) avoiding heavy bleeding, weight changes, libido changes, and non-hormonal; (2) long-acting, avoiding daily dosing, permitting self-discontinuation, and avoiding intermittent bleeding; and (3) concealability without an effectiveness preference. Immediate return to fertility was modeled independently due to lack of clustering. The first dimension was positively correlated with condom use and inversely with implant use. The second was also positively correlated with condom use, and use of both implants and tubal ligation, while inversely with injectable and oral contraceptive pill use. The third was correlated with injectable use.

**Conclusion:** Method attribute preferences do not always align with methods used and may cluster in ways that cannot be satisfied by existing methods.

**Implications:** Preferences for methods that are long-acting, avoid daily dosing, and permit self-discontinuation are widely held and related to method selection, but cannot be met by a single method when held in combination. Client-centered counseling can elicit relative strength of preferences and tailor guidance accordingly, improving method satisfaction.

## 1. INTRODUCTION

In sub-Saharan Africa, nearly one quarter of women of reproductive age have an unmet need for contraception; estimates of unmet need among women living with HIV (WLWH) exceed 40%.[1–3] WLWH have unique contraceptive needs, including concerns about potential interactions between antiretroviral therapy (ART) and contraceptive methods, plans for safe conception, and prevention of vertical HIV transmission. Client-centered counseling can help women select preferred and appropriate family planning (FP) methods and reduce contraceptive discontinuation;[4–7] conversely, lack of incorporation of women’s contraceptive preferences into counseling approaches may lead to unintended pregnancy and other adverse maternal and child health outcomes.

Contraceptive method attributes, including method effectiveness and side effect profiles, should be discussed during counseling; yet women and providers may differ in their perceptions of which method attributes should be prioritized during counseling. As contraceptive preferences are highly individual and complex, tailored counseling is warranted.[8] However, providers may use counseling approaches that focus on specific attributes (such as effectiveness) over others,[9] or hold biases about readiness for pregnancy that impact counseling.[10] Preferences for multiple attributes may also make method selection complex, as some attribute combinations do not align with existing methods. Further, providers offering integrated HIV and reproductive health (RH) care have limited time to provide comprehensive care and limited training to offer contraceptive counseling.[11] While contraceptive preferences and desires can be used to guide client-centered counseling,[12] the degree to which they ultimately impact contraceptive method selection is poorly understood among WLWH.

Preferences for specific method attributes have been shown to align with use of methods with these attributes in some studies, but which attributes align with use and the extent to which they do so varies between studies.[12–14] Additionally, the number and range of attributes studied is limited, and some attributes, including side effect tolerability and concealability, have not been well-characterized. Furthermore, relationships between method attribute preferences and use have not been explored among women in low- and middle-income countries (LMICs), including in sub-Saharan Africa, or among WLWH.

Understanding key contraceptive preference attributes that drive method selection could help improve counseling and increase satisfaction while reducing method switch and discontinuation. We examined alignment of contraceptive method attribute preferences and attributes of methods used among Kenyan WLWH. We also identified clusters of contraceptive method preferences to characterize attributes women often hold in conjunction, and examined associations between clusters and methods used.

## 2. METHODS

### 2.1 Study design, study setting, and study population

We conducted a cross-sectional analysis using baseline survey data from WLWH enrolled in a cluster randomized controlled trial (cRCT), Mobile WACh Empower, evaluating a digital RH counseling and text communication intervention between December 2022 and June 2024.[15] Non-pregnant WLWH of reproductive age (18-45 years; 14-17 if an emancipated minor) who were receiving HIV care at a study site were recruited from 10 HIV clinics, five in Western Kenya and five in Nairobi. Women needed to speak English, Kiswahili, or Luo; have daily access to a mobile phone with a Safaricom SIM; plan to receive HIV care at the study site for the next two years; and be able to read or be comfortable with someone else reading study-related texts. Women participating in another intervention study were ineligible. Women interested in the cRCT were screened for eligibility and provided written consent to enroll. A total of 3,298 women, ∼330 per site, were enrolled and retained for analysis. Additionally, to be included in this analysis, participants must have reported using an eligible contraceptive method (tubal ligation, oral contraception pills [OCPs], injectable, intrauterine device [IUD], implant, or condoms) in the month prior to enrollment. We excluded women who used only traditional methods,[16] vaginal rings, lactational amenorrhea method, or emergency contraception as the sample size was insufficient to characterize method use among these women (n<20).

Ethical approval for this study was granted by the University of Washington’s Human Subjects Division (STUDY00000438) and the Ethics and Research Committee at Kenyatta National Hospital in Kenya (#P162/03/2022).

### 2.2 Data collection

Participants were administered electronic surveys to collect demographic characteristics and RH history. Enrollment was completed at the next scheduled visit, between one week and one year later based on differentiated HIV service delivery schedules in Kenya. At enrollment, all participants completed self-administered, tablet-based surveys on contraceptive attribute preferences. Participants in the intervention arm also received reproductive life counseling on the tablet, shared a report from the summary screen of the counseling tool with HIV care providers, and began receiving RH counseling text communication post-visit.

### 2.3 Contraceptive method attribute and preference definitions

Eleven contraceptive method attribute preferences were captured: preferences for effectiveness, concealability, immediate return to fertility; for methods that are long-acting, avoid daily dosing, permit self-discontinuation; and for methods that avoid intermittent bleeding, heavy bleeding, weight changes, libido changes, and hormones. Preferences were derived from yes/no survey questions, selecting an attribute as one of the three most important, or a combination of the two.(**Table 1**) Preference-use alignment was defined as using a method that satisfies an indicated preference.

**Table 1:**
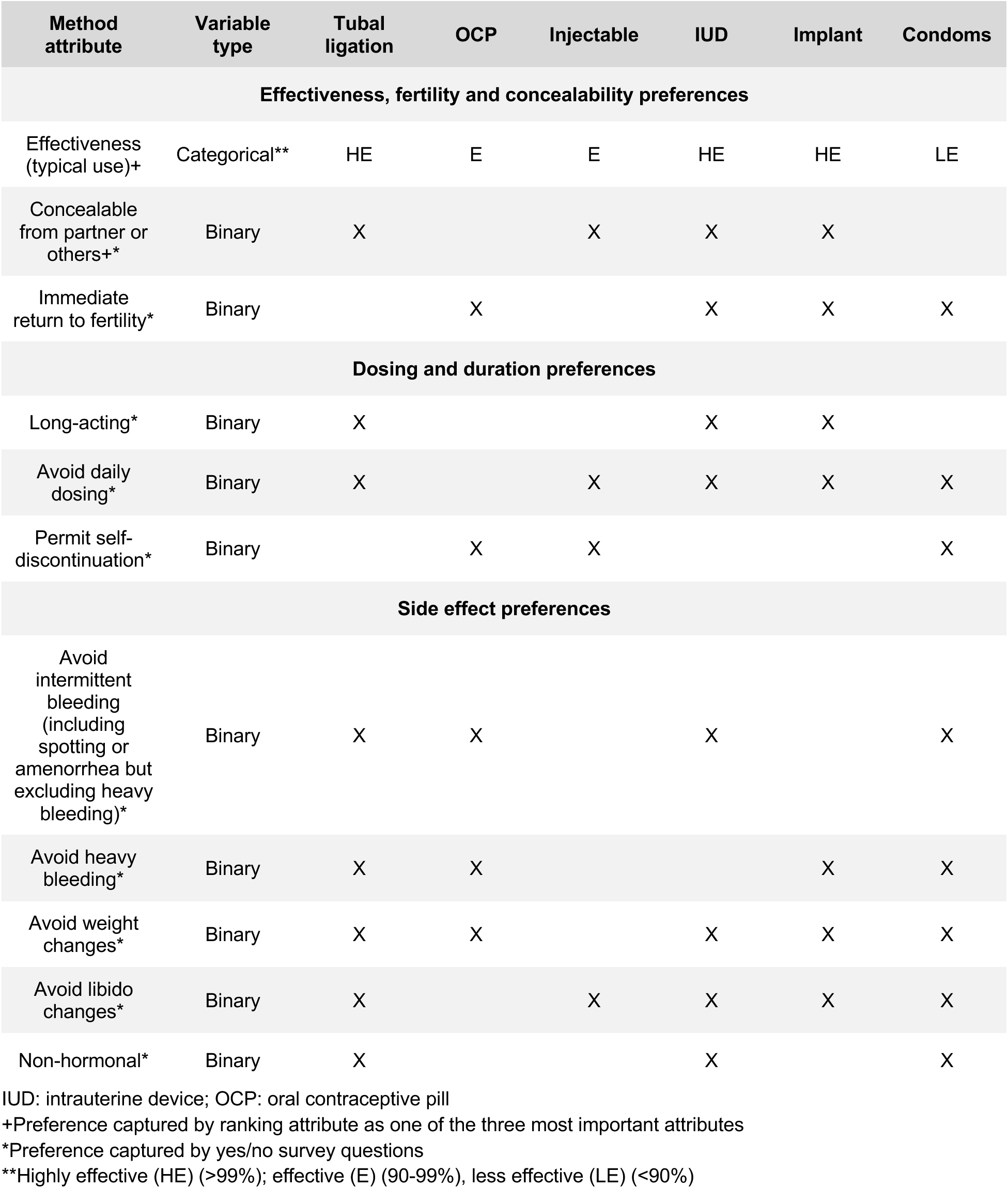
Contraceptive method attributes, by method type.

IUDs, implants, and tubal ligation were classified as highly effective methods (>99% effective with typical use), OCPs and injectables as effective methods (90-99% effective), and male condoms as less effective (<90% effective), in agreement with World Health Organization classifications.(**Table 1**)[16] Other attributes were classified by the research team based on current practice and literature as shown in Table 1.

### 2.4 Data analysis

We compared preferences and method use among women with complete data using a chi-squared test with Donner’s adjustment for clustering. Poisson generalized linear models were constructed with robust standard errors, and accounting for clustering by site, to measure the relationship between each contraceptive attribute preference and use of a method with that attribute to capture preference-use alignment among women with complete data. This “modified Poisson” model structure was used to estimate prevalence ratios (PRs) and is appropriate when the outcome is not rare.[17] Sociodemographic characteristics (age, income, education, marital status) and duration of contraceptive use were assessed as potential covariates for all attribute models; additional covariates were evaluated for inclusion in attribute specific models based on hypothesized relationships between attribute preferences and use. Covariates were individually evaluated in crude models for each attribute, and those with p<0.1 were included in a multivariate model.

In addition, a principal component analysis (PCA) was conducted to identify contraceptive attribute preferences that cluster together to facilitate understanding of factors that influence method selection; women with missing data were excluded from the analysis. This technique reduces the dimension of correlated data into fewer variables that capture as much information from initial variables as possible.[18] Method preferences were included in the initial PCA, and preferences with a uniqueness score >60% (which suggests weak correlation with other preferences) were dropped and modeled separately.[19] Remaining preferences underwent subsequent PCA to generate dimensions, which were retained if the eigenvalue was ≥1 (Kaiser’s criterion).[20] Retained dimensions were transformed using varimax rotation, which aids in interpretation of factor loadings.[18] Dimension loadings with an absolute value ≥0.5 hold the most weight and were used to determine the composition of each dimension, revealing preference clusters.[19] Modified Poisson regression models were used to determine relationships between dimensions and method use, in crude models and multivariate models that included dimensions with p<0.1, accounting for clustering by site. Analyses were conducted using R version 4.4.0.[21]

## 3. RESULTS

Among 3,298 women enrolled and retained in the cRCT, 2,599 (79%) were included in this analysis.(**Figure 1**) Median age was 33 years (interquartile range [IQR] 28-38)(**Table 2**). Most (66%) were married or cohabitating; 58% had three or more living children. The most common contraceptive methods used were implants (36%), injectables (35%), and condoms (34%, including 18% with another method) followed by OCPs (8%), IUDs (4%), and tubal ligation (2%).

**Figure 1:**
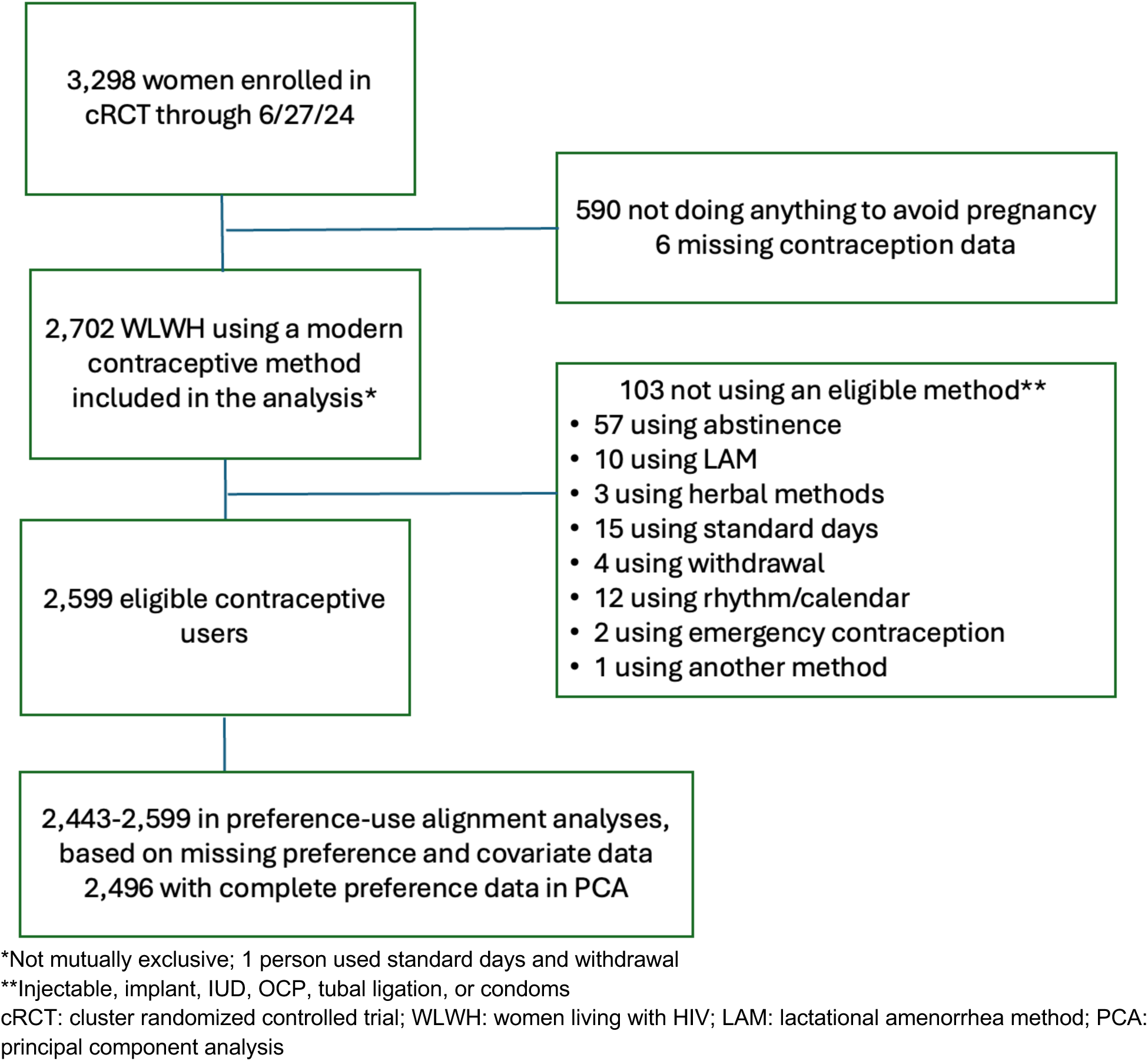
Study flow chart of eligibility for Kenyan WLWH 2022-24

**Table 2:** Characteristics of Kenyan WLWH using contraception and receiving HIV care 2022-24.

| Characteristic | Total<br>N = 2599 | Overall<br>n (%) or median (IQR) |
| --- | --- | --- |
| **Sociodemographic** |  |  |
| Age (years) | 2599 | 33 (28-38) |
| Age category (years) | 2599 |  |
| <25 |  | 288 (11) |
| 25-34 |  | 1174 (45) |
| >34 |  | 1137 (44) |
| Completed at least secondary education | 2599 | 1211 (47) |
| Household income ≥10,000 KSH/month | 2599 | 1144 (44) |
| Employed | 2598 | 1140 (44) |
| Married/cohabitating | 2599 | 1726 (66) |
| Time to clinic (minutes) | 2599 | 30 (30-50) |
| **Clinical characteristics** |  |  |
| Time since HIV diagnosis (years) | 2583 | 8 (4-12) |
| Parity | 2522 | 3 (2-4) |
| Number of living children | 2522 | 3 (2-4) |
| Currently breastfeeding | 2521 | 594 (24) |
| ART use | 2599 | 2579 (99) |
| Efavirenz | 2571 | 37 (1) |
| Dolutegravir | 2578 | 2304 (89) |
| Other | 2566 | 234 (9) |
| **Partner characteristics** |  |  |
| Has regular partner | 2589 | 2007 (77) |
| Partner HIV status | 2007 |  |
| Positive |  | 1100 (55) |
| Negative |  | 681 (34) |
| Don't know |  | 226 (11) |
| Partner on ART | 1095 | 1017 (93) |
| Partner on PrEP | 782 | 286 (37) |
| Disclosed HIV status to partner | 1966 | 1856 (94) |
| **Contraception use and history** |  |  |
| Methods used in prior month | 2599 |  |
| Tubal ligation |  | 52 (2) |
| Implant |  | 933 (36) |
| IUD |  | 93 (4) |
| Injectable |  | 905 (35) |
| OCP |  | 200 (8) |
| Condoms |  | 877 (34) |
| Dual method use in prior month |  | 455 (18) |
| Duration of current method use ≥1 year | 2542 | 1565 (62) |
| Obtained preferred method at most recent visit | 2597 | 2456 (95) |
| Method switch (past year) | 2599 | 177 (7) |
| Discussion of FP with social network in past year | 2588 | 1101 (43) |
| Number of methods used in lifetime | 2599 | 2 (1-3) |

WLWH: women living with HIV; IQR: interquartile range; KSH: Kenyan shilling; ART: antiretroviral therapy; PrEP: pre-exposure prophylaxis; IUD: intrauterine device; OCP: oral contraceptive pills; FP: family planning

### 3.1 Contraceptive method attribute preferences

Overall, the prevalence of preferences for contraceptive method attributes ranged from 85% for avoiding intermittent bleeding to 14% for concealability.(**Figure 2**) The majority of women indicated preferences for methods based on effectiveness (72%) or that are long-acting (60%), non-hormonal (52%), avoid daily dosing (71%), avoid heavy bleeding (63%), or permit self-discontinuation (51%). Preferences for methods that are long-acting, non-hormonal, avoid intermittent bleeding, or highly effective were more prevalent than use of methods with each of these attributes, suggesting unmet preferences.

**Figure 2:**
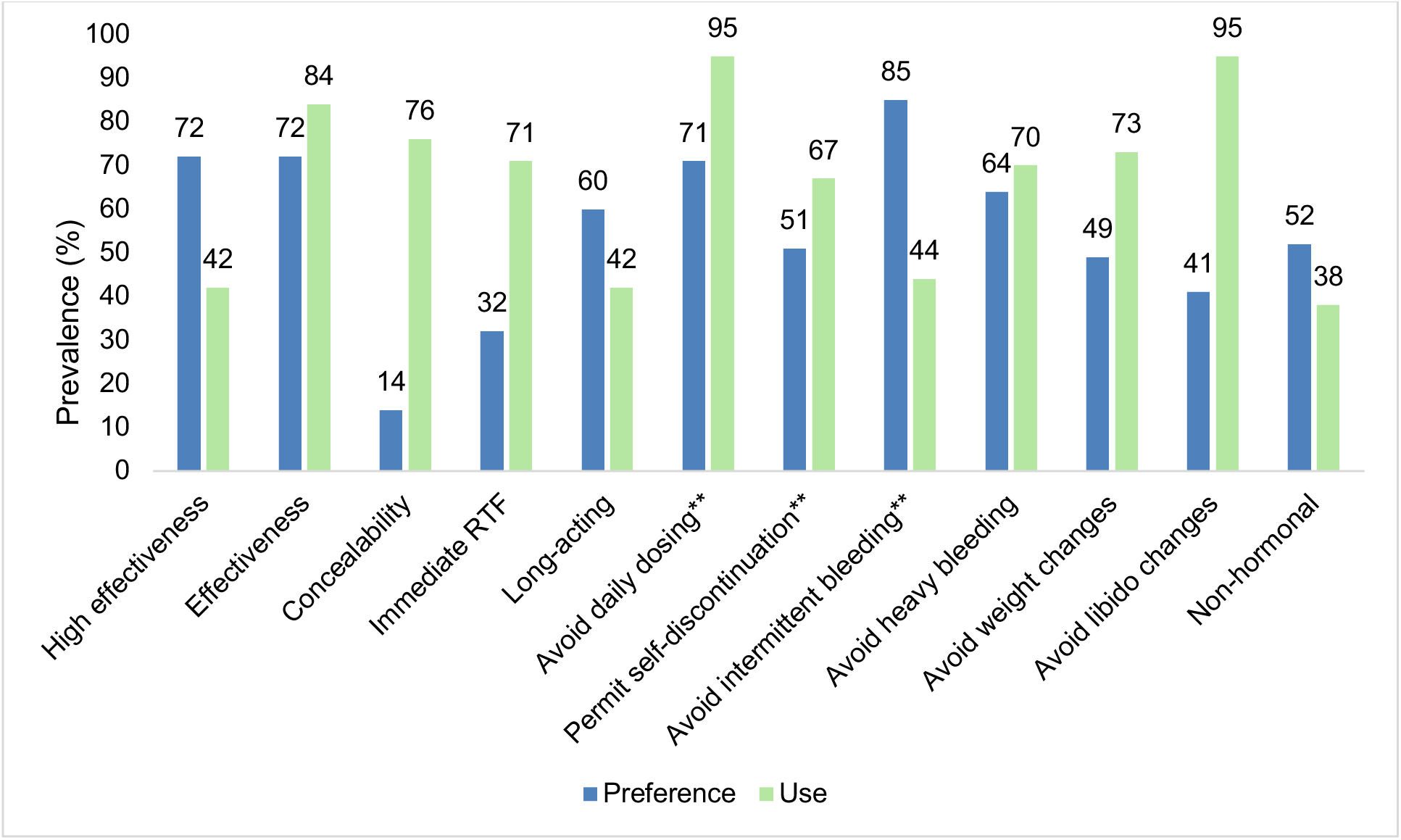
Prevalence of contraceptive method attribute preferences and contraceptive method attributes used among Kenyan WLWH 2022-24 (complete cases, n=2,496)

Several method attributes demonstrated preference-use alignment. Two-thirds (67%) of women who preferred long-acting methods used long-acting methods, compared to 55% of women who did not indicate this preference (aPR:1.63, 95% CI:1.19-2.24)(**Table 3**). Women with preferences for methods that avoided daily dosing were also more likely to use a method other than OCPs (i.e. methods without daily dosing) compared to women without this preference (73% vs. 39%, respectively; aPR:1.11, 95% CI:1.07-1.16). Preferences for methods that permit self-discontinuation were more commonly reported among women who used a method that could be self-discontinued (58%) than among women using a method that required a provider to discontinue (37%)(aPR:1.32, 95% CI:1.14-1.52). Finally, 14% of women preferred concealable methods of contraception which was more common among women who used a concealable method compared to women using a non-concealable method (15% vs. 11%, respectively; aPR: 1.06, 95% CI:1.01-1.12).

**Table 3:** Models of concordance between contraceptive method preferences and method use among Kenyan WLWH 2022-24 (n=2,599)

| Method attribute preferences | Method without corresponding attribute used |  | Method with corresponding attribute used |  | Poisson generalized linear models |  |  |  |
| --- | --- | --- | --- | --- | --- | --- | --- | --- |
|  | N | (%) | N | (%) | Crude PR (95% CI) | p | Adjusted PR (95% CI) | p |
| High effectiveness<sup>a,b,c,d,e,f</sup> | 1097 | (72) | 779 | (72) | 0.97 (0.86, 1.10) | 0.61 | 0.98 (0.88, 1.10) | 0.76 |
| Effectiveness<sup>a,b,c,d,e,g</sup> | 323 | (76) | 1553 | (71) | 0.98 (0.95, 1.02) | 0.44 | 1.00 (0.96, 1.03) | 0.83 |
| Concealability<sup>a,b,c</sup> | 65 | (11) | 297 | (15) | 1.07 (1.01, 1.13) | **0.02** | 1.06 (1.01, 1.12) | **0.03** |
| Immediate return to fertility<sup>a,d</sup> | 238 | (31) | 599 | (33) | 0.99 (0.92, 1.06) | 0.75 | 0.98 (0.92, 1.04) | 0.47 |
| Long-acting<sup>a,b,c,d,e</sup> | 835 | (55) | 719 | (67) | 1.77 (1.28, 2.45) | **<0.001** | 1.63 (1.19, 2.24) | **0.003** |
| Avoid daily dosing<sup>b,c,d</sup> | 56 | (39) | 1769 | (73) | 1.10 (1.06, 1.15) | **<0.001** | 1.11 (1.07, 1.16) | **<0.001** |
| Permits self-discontinuation<sup>b,c,d,f</sup> | 311 | (37) | 1008 | (58) | 1.33 (1.16, 1.52) | **<0.001** | 1.32 (1.14, 1.52) | **<0.001** |
| Avoid intermittent bleeding<sup>a,b,d</sup> | 1186 | (82) | 1014 | (89) | 1.42 (0.96, 2.10) | 0.08 | 1.37 (0.94, 2.01) | 0.10 |
| Avoid heavy bleeding<sup>a,d,h</sup> | 488 | (63) | 1158 | (64) | 1.02 (0.97, 1.07) | 0.51 | 0.99 (0.95, 1.03) | 0.49 |
| Avoid weight changes<sup>a,d</sup> | 323 | (45) | 948 | (50) | 1.04 (0.99, 1.10) | 0.11 | 1.03 (0.97, 1.08) | 0.33 |
| Avoid libido changes<sup>b,c,d</sup> | 44 | (31) | 1015 | (42) | 1.02 (0.99, 1.14) | 0.12 | 1.02 (0.99, 1.05) | 0.16 |
| Non-hormonal<sup>a,b,d</sup> | 777 | (49) | 556 | (56) | 1.15 (0.97, 1.37) | 0.10 | 1.17 (0.99, 1.37) | 0.07 |

PR: prevalence ratio; CI: confidence interval
Adjustment covariates:
a: Age category
b: Income
c: Marital status
d: Contraception use duration
e: Number of children
f: Employment
g: Breastfeeding status
h: Education

We did not detect preference-use alignment for the following method attributes: effectiveness, immediate return to fertility, avoiding intermittent or heavy bleeding, weight changes, libido changes, or hormonal methods.

### 3.2 Method preference clustering

#### 3.2.1 PCA analysis

In analysis of 2,496 participants with complete method preference data, preference for immediate return to fertility had a uniqueness score of 0.69 and was excluded from subsequent PCAs and modeled separately. We identified three dimensions with eigenvalues >1, explaining 58% of total variance. Factor loadings with an absolute value of ≥0.5 for each dimension included preferences for methods that: Dimension 1) avoid heavy bleeding, weight changes, and libido changes, as well as for non-hormonal methods [side-effect related preferences]; Dimension 2) are long-acting, avoid daily dosing, permit self-discontinuation, and avoid intermittent bleeding [dosing, duration, and regular menses preferences]; and Dimension 3) are concealable but no preference for effectiveness [concealability but not effectiveness preferences] (**Table 4**).

**Table 4:**
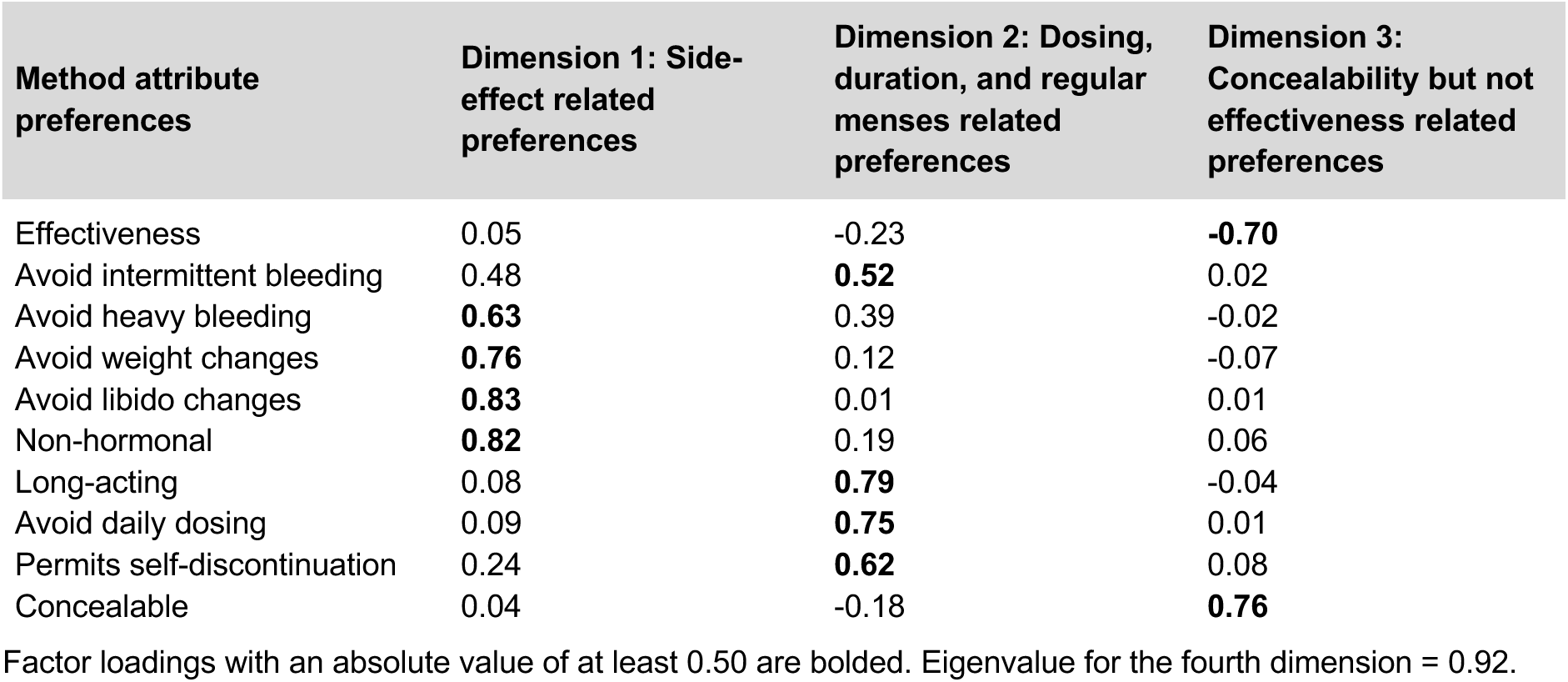
Varimax-rotated loading matrix of Kenyan WLWH’s contraceptive attribute preferences 2022-24.

| Method attribute preferences | Dimension 1: Side-effect related preferences | Dimension 2: Dosing, duration, and regular menses related preferences | Dimension 3: Concealability but not effectiveness related preferences |
| --- | --- | --- | --- |
| Effectiveness | 0.05 | -0.23 | **-0.70** |
| Avoid intermittent bleeding | 0.48 | **0.52** | 0.02 |
| Avoid heavy bleeding | **0.63** | 0.39 | -0.02 |
| Avoid weight changes | **0.76** | 0.12 | -0.07 |
| Avoid libido changes | **0.83** | 0.01 | 0.01 |
| Non-hormonal | **0.82** | 0.19 | 0.06 |
| Long-acting | 0.08 | **0.79** | -0.04 |
| Avoid daily dosing | 0.09 | **0.75** | 0.01 |
| Permits self-discontinuation | 0.24 | **0.62** | 0.08 |
| Concealable | 0.04 | -0.18 | **0.76** |

Factor loadings with an absolute value of at least 0.50 are bolded. Eigenvalue for the fourth dimension = 0.92.

#### 3.2.3 Poisson models

Women with side-effect related preferences (Dimension 1) were more likely to use condoms (aPR:1.11, 95% CI:1.02-1.20), but less likely to use implants (aPR:0.91, 95% CI:0.85-0.97)(**Table 5**). Women with dosing, duration, and regular menses-related preferences, Dimension 2, were similarly more likely to use condoms (aPR:1.13, 95% CI:1.02-1.26), as well as implants (aPR:1.12, 95% CI:1.04-1.20), IUDs (aPR:1.17, 95% CI:1.04-1.32) and tubal ligation (aPR:1.48, 95% CI:1.08-2.02), but less likely to use injectables (aPR:0.92, 95% CI: 0.86-0.98) and OCPs (aPR:0.72, 95% CI:0.61-0.85). In contrast, injectable use was higher among women with concealability but not effectiveness-related preferences, Dimension 3 (aPR:1.09, 95% CI:1.03-1.16).

**Table 5:** Correlations between PCA dimensions and method use among Kenyan WLWH 2022-24.

| PCA Dimension | Poisson generalized linear models |  |  |  |
| --- | --- | --- | --- | --- |
|  | Crude PR (95% CI) | p | Adjusted PR (95% CI) | p |
| <u>Condom use</u> |  |  |  |  |
| Dimension 1* | 1.09 (1.00, 1.19) | **0.04** | 1.11 (1.02, 1.20) | **0.01** |
| Dimension 2** | 1.12 (0.99, 1.26) | **0.07** | 1.13 (1.02, 1.26) | **0.03** |
| Dimension 3*** | 1.00 (0.95, 1.05) | 0.92 | -- | -- |
| Immediate RTF preference | 1.08 (0.93, 1.27) | 0.30 | -- | -- |
| <u>Implant use</u> |  |  |  |  |
| Dimension 1 | 0.90 (0.84, 0.96) | **0.003** | 0.91 (0.85, 0.97) | **0.005** |
| Dimension 2 | 1.13 (1.03, 1.24) | **0.01** | 1.12 (1.04, 1.20) | **0.003** |
| Dimension 3 | 0.95 (0.88, 1.02) | 0.19 | -- | -- |
| Immediate RTF preference | 0.96 (0.87, 1.06) | 0.39 | -- | -- |
| <u>IUD use</u> |  |  |  |  |
| Dimension 1 | 0.89 (0.65, 1.22) | 0.47 | -- | -- |
| Dimension 2 | 1.16 (1.02, 1.31) | **0.02** | 1.17 (1.04, 1.32) | **0.009** |
| Dimension 3 | 1.16 (0.97, 1.39) | **0.10** | 1.17 (0.98, 1.40) | 0.08 |
| Immediate RTF preference | 0.74 (0.47, 1.17) | 0.20 | -- | -- |
| <u>Injectable use</u> |  |  |  |  |
| Dimension 1 | 1.04 (0.98, 1.10) | 0.23 | -- | -- |
| Dimension 2 | 0.91 (0.85, 0.98) | **0.01** | 0.92 (0.86, 0.98) | **0.01** |
| Dimension 3 | 1.10 (1.04, 1.17) | **0.002** | 1.09 (1.03, 1.16) | **0.004** |
| Immediate RTF preference | 1.07 (0.94, 1.22) | 0.29 | -- | -- |
| <u>OCP use</u> |  |  |  |  |
| Dimension 1 | 0.94 (0.79, 1.12) | 0.51 | -- | -- |
| Dimension 2 | 0.71 (0.61, 0.82) | **<0.001** | 0.72 (0.61, 0.85) | **<0.001** |
| Dimension 3 | 0.88 (0.75, 1.04) | 0.14 | -- | -- |
| Immediate RTF preference | 0.72 (0.49, 1.06) | **0.10** | 0.81 (0.54, 1.22) | 0.31 |
| <u>Tubal ligation use</u> |  |  |  |  |
| Dimension 1 | 0.73 (0.51, 1.04) | **0.09** | 0.87 (0.55, 1.36) | 0.54 |
| Dimension 2 | 1.48 (1.06, 2.07) | **0.02** | 1.48 (1.08, 2.02) | **0.01** |
| Dimension 3 | 0.89 (0.75, 1.06) | 0.20 | -- | -- |
| Immediate RTF preference | 0.38 (0.21, 0.71) | **0.002** | 0.38 (0.17, 0.88) | **0.02** |

\*Dimension 1: Side-effect related preferences
\*\*Dimension 2: Dosing, duration, and regular menses preferences
\*\*\*Dimension 3: Concealability but not effectiveness preferences
PCA: principal component analysis; PR: prevalence ratio; CI: confidence interval; RTF: return to fertility; IUD: intrauterine device; OCP: oral contraceptive pills

## 4. DISCUSSION

In a cohort of Kenyan WLWH using modern contraception, most preferred contraceptive methods based on their effectiveness, or which are long-acting, avoid daily dosing, or avoid bleeding-related side effects, with at least 60% of women desiring each of these method attributes. Preferences aligned with use of methods that were concealable, long-acting, avoid daily dosing, and permit self-discontinuation.

While nearly three-quarters of women in our study indicated a preference for effectiveness, only 41% of women used a highly effective method; these results concur with other studies.[12,13] Women may select methods with lower effectiveness because other method attributes are determined to be more important. Alternatively, women may perceive or define effectiveness differently than clinical standards of effectiveness, and/or may not require the highest level of effectiveness to be comfortable selecting a method.[13] Feelings about effectiveness may also vary based on the strength of desire for pregnancy prevention, or ambivalent feelings about pregnancy.

In our study, we found that convenience-related preferences (dosing, duration of use, and ability to self-discontinue) were co-occurring, common, and associated with using methods with these attributes. A systematic review among people living with HIV similarly found that method convenience was frequently cited among preferences for contraception.[22] The burden of remembering a daily pill, which may be seen as inconvenient, was a deterrent among WLWH and was associated with low use of OCPs in this population.[23] Furthermore, WLWH have been shown to be more than five times more likely to use OCPs prior to HIV diagnosis as opposed to following diagnosis, suggesting pills may be unfavorable due to the added pill burden or stigma associated with pills used for HIV treatment. The relationship we detected between the convenience dimension and condom use also aligns with prior studies; accessibility and convenience are frequently cited as the rationale for using male condoms in sub-Saharan Africa.[22]

We found that preferences related to method side effects clustered together and were correlated with condom use, the only non-permanent contraceptive method in this analysis with minimal potential side effects. Separately, we found preference-use alignment for concealability.

In PCA, a preference for concealability clustered with the lack of preference for effectiveness, suggesting desire for concealability may override other preferences and drive method selection.

Many women expressed preferences for methods that are long-acting, avoid daily dosing, and are highly effective, attributes consistent with existing long-acting reversible contraceptive (LARC) methods; however, these preferences did not cluster in a single dimension. In contrast, convenience-related preferences for methods that were long-acting, avoid daily dosing, and permit self-discontinuation, which did cluster together, do not typically correspond with existing methods (except IUD self-removal, which has not been well-studied in LMICs).[24] Thus, women are often forced to make tradeoffs when selecting a method. This supports the need for innovation in contraceptive development, including methods that meet unique preference combinations and novel methods that can be self-discontinued. When women must choose between methods with different attribute combinations, client-centered counseling is essential to elicit preferences and tailor counseling accordingly to guide women through the process of method selection to maximize satisfaction and autonomy.

Our study has several strengths. We assessed a diverse array of preferences, permitting a complex analysis of the strength of contraceptive attribute preference-use alignment in a large cohort of WLWH, whose preferences for contraception have been understudied. Furthermore, we used a robust analytical approach to identify how attribute preferences co-occur.

Our study is also subject to limitations. We were unable to assess the temporal relationship between contraceptive method preferences and methods used due to the cross-sectional study design. In addition, our results may not be generalizable to WLWH from other regions or countries. We did not examine preferences for those with potential unmet need for contraception, which could lead to selection bias. Contraceptive attribute preferences were captured in multiple ways and bias could be introduced by capturing preferences with survey questions that used different formats. Finally, we did not collect data on other potentially important preferences, including the ability to protect against HIV/STI transmission, or the relative ranking/strength of preferences for specific attributes.

In conclusion, Kenyan WLWH identified preferences for methods that are long-acting, avoid daily dosing, and permit self-discontinuation in method selection. WLWH hold complex contraceptive preferences that may not be available within a single method. Additional research that aims to understand how contraceptive method attribute preferences inform method selection could help improve client-centered counseling tailored to unique values and preferences of WLWH, and method satisfaction, reducing unintended pregnancies and adverse maternal and child health outcomes.

## Data Availability

Due to the sensitive nature of the data collected for this study, access is restricted. Requests for access to anonymized data may be made to the corresponding author and will be considered on a case-by-case basis, subject to ethical approval.

## Acknowledgements and funding

We would like to acknowledge the contributions from study participants and staff. This study was made possible through funding and support from NIH/NICHD R01HD104551, the University of Washington/Fred Hutch Center for AIDS Research (CFAR) (NIH/NIAID P30 AI027757), and the University of Washingtons’ Global Center for Integrated Health of Women, Adolescents, and Children (Global WACh).

## Notes

### Competing Interest Statement

The authors have declared no competing interest.

### Author Declarations

The Human Subjects Division of University of Washington gave ethical approval for this work (STUDY00000438), and the Ethics and Research Committee at Kenyatta National Hospital in Kenya (#P162/03/2022) gave ethical approval for this work.

