## Supplemental Tables for "Alignment of contraceptive method attribute preferences and use among Kenyan women living with HIV"

Table 3a: Concordance between preference for and use of highly effective method

| Method attribute preferences and potential confounders | Non-highly effective method used (N=1522) | | Highly effective method used (N=1077) | | Poisson generalized linear models | | | |
|---|---|---|---|---|---|---|---|---|
| | **N** | **(%)** | **N** | **(%)** | **Crude PR (95% CI)** | **p** | **Adjusted PR (95% CI)** | **p** |
| *Method attribute preference* | | | | | | | | |
| Effectiveness | 1097 | (72) | 779 | (72) | 1.01 (0.89, 1.16) | 0.88 | 1.03 (0.91, 1.19) | 0.62 |
| Concealability | 217 | (14) | 145 | (13) | 0.96 (0.80, 1.14) | 0.66 | -- | -- |
| *Demographic characteristics* | | | | | | | | |
| Age | | | | | | | | |
| <25 | 152 | (10) | 136 | (13) | Ref. | Ref. | Ref. | Ref. |
| 25-34 | 668 | (44) | 506 | (47) | 0.91 (0.76, 1.11) | 0.34 | 0.72 (0.59, 0.89) | **0.002** |
| >34 | 702 | (46) | 435 | (40) | 0.81 (0.67, 0.99) | **0.03** | 0.58 (0.47, 0.73) | **<0.001** |
| Income ≥10,000 KSH/month | 712 | (47) | 432 | (40) | 0.85 (0.75, 0.96) | **0.01** | 0.87 (0.77, 0.99) | **0.03** |
| At least secondary education | 739 | (49) | 472 | (44) | 0.89 (0.79, 1.01) | **0.07** | 0.96 (0.84, 1.10) | 0.55 |
| Married/cohabitating | 957 | (63) | 769 | (71) | 1.26 (1.11, 1.44) | **<0.001** | 1.18 (1.03, 1.35) | **0.02** |
| Contraception use ≥1 year | 829 | (54) | 736 | (68) | 1.35 (1.19, 1.54) | **<0.001** | 1.46 (1.28, 1.67) | **<0.001** |
| Number of children | 2.9 | (1.4) | 3.0 | (1.4) | 1.04 (1.00, 1.09) | **0.06** | 1.07 (1.01, 1.12) | **0.01** |
| Currently breastfeeding | 332 | (22) | 262 | (24) | 1.07 (0.93, 1.22) | 0.35 | -- | -- |
| Employed | 681 | (45) | 459 | (43) | 0.95 (0.84, 1.07) | 0.37 | -- | -- |

Table 3b: Concordance between preference for and use of effective method

| Method attribute preferences and potential confounders | Less effective method use (N=423) | | Effective method use (N=2176) | | Poisson generalized linear models | | | |
|---|---|---|---|---|---|---|---|---|
| | N | (%) | N | (%) | Crude PR (95% CI) | p | Adjusted PR (95% CI) | p |
| *Method attribute preference* | | | | | | | | |
| Effectiveness | 323 | (76) | 1553 | (71) | 0.96 (0.88, 1.05) | 0.40 | 0.98 (0.89, 1.08) | 0.67 |
| *Demographic characteristics* | | | | | | | | |
| Age | | | | | | | | |
| <25 | 52 | (12) | 236 | (11) | Ref. | Ref. | -- | -- |
| 25-34 | 142 | (34) | 1032 | (47) | 1.07 (0.93, 1.24) | 0.33 | -- | -- |
| >34 | 229 | (54) | 908 | (42) | 0.97 (0.85, 1.13) | 0.72 | -- | -- |
| Income ≥10,000 KSH/month | 226 | (53) | 918 | (42) | 0.93 (0.85, 1.01) | **0.09** | 0.95 (0.87, 1.03) | 0.21 |
| At least secondary education | 209 | (49) | 1002 | (46) | 0.98 (0.90, 1.06) | 0.61 | -- | -- |
| Married/cohabitating | 221 | (52) | 1505 | (69) | 1.13 (1.04, 1.24) | **0.007** | 1.10 (1.00, 1.20) | **0.05** |
| Contraception use ≥1 year | 276 | (74) | 1289 | (59) | 0.92 (0.84, 1.00) | **0.04** | 0.92 (0.85, 1.01) | 0.07 |
| Number of children | 2.7 | (1.5) | 3.0 | (1.4) | 1.02 (0.99, 1.05) | 0.18 | -- | -- |
| Currently breastfeeding | 67 | (18) | 527 | (25) | 1.06 (0.97, 1.17) | 0.21 | -- | -- |
| Employed | 196 | (46) | 944 | (43) | 0.98 (0.90, 1.07) | 0.64 | -- | -- |

Table 3c: Concordance between preference for and use of concealable method

| Method attribute preferences and potential confounders | Non-concealable method used (n=618) | | Concealable method used (n=1981) | | Poisson generalized linear models | | | |
|---|---|---|---|---|---|---|---|---|
| | N | (%) | N | (%) | Crude PR (95% CI) | p | Adjusted PR (95% CI) | p |
| *Method attribute preference* | | | | | | | | |
| Concealability | 65 | (11) | 297 | (15) | 1.09 (0.96, 1.23) | 0.17 | 1.09 (0.96, 1.23) | 0.19 |
| *Demographic characteristics* | | | | | | | | |
| Age | | | | | | | | |
| <25 | 71 | (11) | 217 | (11) | Ref. | Ref. | -- | -- |
| 25-34 | 222 | (36) | 952 | (48) | 1.08 (0.93, 1.25) | 0.33 | -- | -- |
| >34 | 325 | (53) | 812 | (41) | 0.95 (0.82, 1.10) | 0.48 | -- | -- |
| Income ≥10,000 KSH/month | 325 | (53) | 819 | (41) | 0.90 (0.82, 0.98) | **0.02** | 0.89 (0.81, 0.97) | **0.009** |
| At least secondary education | 304 | (49) | 907 | (46) | 0.97 (0.89, 1.06) | 0.47 | -- | -- |
| Married/cohabitating | 363 | (59) | 1363 | (69) | 1.12 (1.01, 1.23) | **0.02** | 1.12 (1.02, 1.23) | **0.02** |
| Contraception use ≥1 year | 350 | (57) | 1215 | (62) | 1.00 (0.91, 1.09) | 0.97 | -- | -- |
| Discussed FP* with someone other than HCW* in prior year | 265 | (43) | 836 | (42) | 1.00 (0.91, 1.09) | 0.92 | -- | -- |
| Disclosure of HIV status to partner+ | 426 | (93) | 1430 | (95) | 1.07 (0.86, 1.36) | 0.54 | -- | -- |

*FP: Family planning; HCW: Health care worker

+ among those with a regular partner

Table 3d: Concordance between preference for and use of method with immediate return to fertility

| Method attribute preferences and potential confounders | Use of method with delayed RTF* (N=758) | | Use of method with immediate RTF* (N=1834) | | Poisson generalized linear models | | | |
|---|---|---|---|---|---|---|---|---|
| | N | (%) | N | (%) | Crude PR (95% CI) | p | Adjusted PR (95% CI) | p |
| *Method attribute preference* | | | | | | | | |
| Model 1: Immediate return to fertility | 238 | (31) | 599 | (33) | 1.02 (0.92, 1.12) | 0.74 | 1.01 (0.91, 1.11) | 0.88 |
| *Demographic characteristics* | | | | | | | | |
| Age | | | | | | | | |
| <25 | 67 | (9) | 220 | (12) | Ref. | | -- | -- |
| 25-34 | 361 | (48) | 811 | (44) | 0.91 (0.78, 1.05) | 0.20 | -- | -- |
| >34 | 330 | (44) | 803 | (44) | 0.93 (0.80, 1.08) | 0.33 | -- | -- |
| Income ≥10,000 KSH/month | 320 | (42) | 818 | (45) | 1.03 (0.94, 1.13) | 0.56 | -- | -- |
| At least secondary education | 348 | (46) | 860 | (47) | 1.01 (0.92, 1.11) | 0.85 | -- | -- |
| Married/cohabitating | 496 | (65) | 1225 | (67) | 1.02 (0.93, 1.12) | 0.70 | -- | -- |
| Contraception use ≥1 year | 402 | (53) | 1158 | (65) | 1.17 (1.06, 1.29) | **0.002** | 1.16 (1.05, 1.28) | **0.003** |
| Number of children | 3.0 | (1.4) | 2.9 | (1.4) | 0.97 (0.94, 1.01) | 0.12 | -- | -- |
| Currently breastfeeding | 170 | (23) | 424 | (24) | 1.02 (0.91, 1.13) | 0.74 | -- | -- |

*RTF= Return to fertility

Table 3e: Concordance between preference for and use of long-acting method

| **Method attribute preferences and potential confounders** | **Short-acting method used (n=1508)** | | **Long-acting method used (n=1069)** | | **Poisson generalized linear models** | | | |
|---|---|---|---|---|---|---|---|---|
| | **N** | **(%)** | **N** | **(%)** | **Crude PR (95% CI)** | **p** | **Adjusted PR (95% CI)** | **p** |
| *Method attribute preference* | | | | | | | | |
| Long-acting | 835 | (55) | 719 | (67) | 1.35 (1.19, 1.54) | **<0.001** | 1.35 (1.17, 1.57) | **<0.001** |
| Avoid daily dosing | 1032 | (68) | 782 | (73) | 1.15 (1.00, 1.32) | **0.05** | 1.00 (0.86, 1.17) | 0.99 |
| *Demographic characteristics* | | | | | | | | |
| Age | | | | | | | | |
| <25 | 150 | (10) | 134 | (13) | Ref. | Ref. | Ref. | Ref. |
| 25-34 | 664 | (44) | 504 | (47) | 0.91 (0.76, 1.11) | 0.34 | 0.73 (0.60, 0.90) | **0.003** |
| >34 | 694 | (46) | 431 | (40) | 0.81 (0.67, 0.99) | **0.03** | 0.59 (0.47, 0.74) | **<0.001** |
| Income ≥10,000 KSH/month | 701 | (47) | 429 | (40) | 0.85 (0.75, 0.96) | **0.01** | 0.85 (0.75, 0.96) | **0.01** |
| At least secondary education | 732 | (49) | 467 | (44) | 0.89 (0.79, 1.01) | **0.07** | 0.96 (0.84, 1.09) | 0.53 |
| Married/cohabitating | 951 | (63) | 761 | (71) | 1.26 (1.11, 1.44) | **<0.001** | 1.17 (1.02, 1.34) | **0.03** |
| Contraception use ≥1 year | 823 | (57) | 730 | (68) | 1.35 (1.19, 1.54) | **<0.001** | 1.44 (1.26, 1.64) | **<0.001** |
| Number of children | 2.9 | (1.4) | 3.0 | (1.4) | 1.04 (1.00, 1.09) | **0.06** | 1.07 (1.02, 1.12) | **0.01** |
| Currently breastfeeding | 332 | (23) | 261 | (25) | 1.07 (0.93, 1.22) | 0.36 | -- | -- |

Table 3f: Concordance between preference for and use of method that avoids daily dosing

| Method attribute preferences and potential confounders | Daily dosing required (n=144) | | Avoids daily dosing (n=2431) | | Poisson generalized linear models | | | |
|---|---|---|---|---|---|---|---|---|
| | N | (%) | N | (%) | Crude PR (95% CI) | p | Adjusted PR (95% CI) | p |
| *Method attribute preference* | | | | | | | | |
| Avoid daily dosing | 56 | (39) | 1769 | (73) | 1.10 (1.00, 1.20) | **0.04** | 1.10 (1.00, 1.20) | **0.04** |
| Long-acting method | 68 | (47) | 1477 | (61) | 1.03 (0.95, 1.12) | 0.48 | -- | -- |
| *Demographic characteristics* | | | | | | | | |
| Age | | | | | | | | |
| <25 | 13 | (9) | 274 | (11) | Ref. | Ref. | -- | -- |
| 25-34 | 59 | (41) | 1103 | (45) | 1.00 (0.88, 1.14) | 0.98 | -- | -- |
| >34 | 72 | (50) | 1054 | (43) | 0.98 (0.86, 1.13) | 0.82 | -- | -- |
| Income ≥10,000 KSH/month | 74 | (51) | 1051 | (43) | 0.98 (0.91, 1.06) | 0.65 | -- | -- |
| At least secondary education | 71 | (49) | 1126 | (46) | 0.99 (0.92, 1.07) | 0.86 | -- | -- |
| Married/cohabitating | 105 | (73) | 1605 | (66) | 0.98 (0.90, 1.07) | 0.68 | -- | -- |
| Contraception use ≥1 year | 52 | (37) | 1498 | (63) | 1.07 (0.98, 1.16) | 0.13 | -- | -- |

Table 3g: Concordance between preference for and use of method that permits self-discontinuation

| Method attribute preferences and potential confounders | Use of method that cannot be self-discontinued (n=843) | | Use of method that can be self-discontinued (n=1724) | | Poisson generalized linear models | | | |
|---|---|---|---|---|---|---|---|---|
| | N | (%) | N | (%) | Crude PR (95% CI) | p | Adjusted PR (95% CI) | p |
| *Method attribute preference* | | | | | | | | |
| Permits self-discontinuation | 311 | (37) | 1008 | (58) | 1.33 (1.21, 1.47) | **<0.001** | 1.32 (1.20, 1.46) | **<0.001** |
| Immediate return to fertility | 248 | (29) | 580 | (34) | 1.06 (0.96, 1.18) | 0.22 | -- | -- |
| *Demographic characteristics* | | | | | | | | |
| Age | | | | | | | | |
| <25 | 100 | (12) | 186 | (11) | Ref. | Ref. | -- | -- |
| 25-34 | 409 | (49) | 749 | (43) | 0.99 (0.84, 1.16) | 0.88 | -- | -- |
| >34 | 334 | (40) | 789 | (46) | 1.07 (0.92, 1.26) | 0.40 | -- | -- |
| Income ≥10,000 KSH/month | 329 | (39) | 799 | (46) | 1.10 (1.00, 1.21) | **0.04** | 1.07 (0.97, 1.18) | 0.16 |
| At least secondary education | 374 | (44) | 826 | (48) | 1.05 (0.96, 1.16) | 0.29 | -- | -- |
| Married/cohabitating | 593 | (70) | 1116 | (65) | 0.92 (0.83, 1.02) | **0.10** | 0.92 (0.83, 1.02) | 0.10 |
| Contraception use ≥1 year | 564 | (67) | 983 | (59) | 0.89 (0.81, 0.98) | **0.02** | 0.89 (0.81, 0.98) | **0.02** |
| Employed | 354 | (42) | 770 | (45) | 1.04 (0.95, 1.14) | 0.41 | -- | -- |

Table 3h: Concordance between preference for and use of method that avoids intermittent bleeding

| **Method attribute preferences and potential confounders** | **Use of method that may cause spotting/amenorrhea (n=1453)** | | **Use of method that avoids intermittent bleeding (n=1142)** | | **Poisson generalized linear models** | | | |
|---|---|---|---|---|---|---|---|---|
| | **N** | **(%)** | **N** | **(%)** | **Crude PR (95% CI)** | **p** | **Adjusted PR (95% CI)** | **p** |
| *Method attribute preference* | | | | | | | | |
| Avoid intermittent bleeding | 1186 | (82) | 1014 | (89) | 1.42 (1.19, 1.72) | **<0.001** | 1.34 (1.08, 1.66) | **0.008** |
| Avoid heavy bleeding | 880 | (61) | 765 | (67) | 1.17 (1.04, 1.33) | **0.01** | 1.04 (0.90, 1.20) | 0.62 |
| *Demographic characteristics* | | | | | | | | |
| Age | | | | | | | | |
| <25 | 163 | (11) | 125 | (11) | Ref. | Ref. | Ref. | Ref. |
| 25-34 | 737 | (51) | 435 | (38) | 0.85 (0.70. 1.05) | 0.12 | 0.87 (0.71, 1.07) | 0.19 |
| >34 | 553 | (38) | 582 | (51) | 1.18 (0.98, 1.44) | **0.09** | 1.18 (0.97, 1.45) | 0.11 |
| Income ≥10,000 KSH/month | 575 | (40) | 566 | (50) | 1.25 (1.11, 1.41) | **<0.001** | 1.21 (1.08, 1.37) | **0.002** |
| At least secondary education | 661 | (45) | 547 | (48) | 1.06 (0.94, 1.19) | 0.36 | -- | -- |
| Married/cohabitating | 987 | (68) | 738 | (65) | 0.92 (0.82, 1.04) | 0.19 | -- | -- |
| Contraception use ≥1 year | 853 | (59) | 712 | (62) | 1.17 (1.04, 1.33) | **0.01** | 1.10 (0.97, 1.25) | 0.13 |

Table 3i: Concordance between preference for and use of method that avoids heavy bleeding

| Method attribute preferences and potential confounders | Use of method that may cause heavy bleeding (n=779) | | Use of method that avoids heavy bleeding (n=1814) | | Poisson generalized linear models | | | |
|---|---|---|---|---|---|---|---|---|
| | **N** | **(%)** | **N** | **(%)** | **Crude PR (95% CI)** | **p** | **Adjusted PR (95% CI)** | **p** |
| *Method attribute preference* | | | | | | | | |
| Avoid heavy bleeding | 488 | (63) | 1158 | (64) | 1.02 (0.92, 1.12) | 0.74 | 1.00 (0.91, 1.10) | 0.98 |
| Avoid intermittent bleeding | 645 | (83) | 1552 | (86) | 1.06 (0.94, 1.22) | 0.35 | -- | -- |
| *Demographic characteristics* | | | | | | | | |
| Age | | | | | | | | |
| <25 | 69 | (9) | 218 | (12) | Ref. | Ref. | -- | -- |
| 25-34 | 378 | (49) | 794 | (44) | 0.89 (0.77, 1.04) | 0.15 | -- | -- |
| >34 | 332 | (43) | 802 | (44) | 0.93 (0.81, 1.09) | 0.37 | -- | -- |
| Income ≥10,000 KSH/month | 336 | (43) | 804 | (44) | 1.01 (0.92. 1.11) | 0.77 | -- | -- |
| At least secondary education | 382 | (49) | 825 | (46) | 0.96 (0.87, 1.05) | 0.36 | -- | -- |
| Married/cohabitating | 515 | (66) | 1208 | (67) | 1.01 (0.92, 1.11) | 0.87 | -- | -- |
| Contraception use ≥1 year | 411 | (53) | 1152 | (65) | 1.18 (1.07, 1.30) | **<0.001** | 1.18 (1.07, 1.30) | **0.001** |

Table 3j: Concordance between preference for and use of method that avoids weight changes

| Method attribute preferences and potential confounders | Use of method that may cause weight changes (n=710) | | Use of method that avoids weight changes (n=1880) | | Poisson generalized linear models | | | |
|---|---|---|---|---|---|---|---|---|
| | **N** | **(%)** | **N** | **(%)** | **Crude PR (95% CI)** | **p** | **Adjusted PR (95% CI)** | **p** |
| *Method attribute preference* | | | | | | | | |
| Avoid weight changes | 323 | (45) | 948 | (50) | 1.06 (0.96, 1.16) | 0.24 | 1.03 (0.94, 1.13) | 0.48 |
| Non-hormonal | 357 | (50) | 974 | (52) | 1.02 (0.93, 1.11) | 0.73 | -- | -- |
| *Demographic characteristics* | | | | | | | | |
| Age | | | | | | | | |
| <25 | 67 | (9) | 220 | (12) | Ref. | Ref. | -- | -- |
| 25-34 | 354 | (50) | 816 | (43) | 0.91 (0.79, 1.06) | 0.22 | -- | -- |
| >34 | 289 | (41) | 844 | (45) | 0.97 (0.84, 1.13) | 0.67 | -- | -- |
| Income ≥10,000 KSH/month | 297 | (42) | 840 | (45) | 1.03 (0.94, 1.13) | 0.52 | -- | -- |
| At least secondary education | 339 | (48) | 869 | (46) | 0.98 (0.90, 1.08) | 0.72 | -- | -- |
| Married/cohabitating | 465 | (65) | 1257 | (67) | 1.02 (0.93, 1.12) | 0.71 | -- | -- |
| Contraception use ≥1 year | 364 | (51) | 1196 | (66) | 1.19 (1.08, 1.31) | **<0.001** | 1.18 (1.07, 1.30) | **<0.001** |

Table 3k: Concordance between preference for and use of method that avoids libido changes

| Method attribute preferences and potential confounders | Use of method that may cause libido changes (n=141) | | Use of method that avoids libido changes (n=2433) | | Poisson generalized linear models | | | |
|---|---|---|---|---|---|---|---|---|
| | N | (%) | N | (%) | Crude PR (95% CI) | p | Adjusted PR (95% CI) | p |
| *Method attribute preference* | | | | | | | | |
| Avoid libido changes | 44 | (31) | 1015 | (42) | 1.02 (0.94, 1.11) | 0.56 | 1.02 (0.94, 1.11) | 0.56 |
| Non-hormonal | 63 | (45) | 1265 | (52) | 1.02 (0.94, 1.10) | 0.67 | -- | -- |
| *Demographic characteristics* | | | | | | | | |
| Age | | | | | | | | |
| <25 | 13 | (9) | 271 | (11) | Ref. | Ref. | -- | -- |
| 25-34 | 56 | (40) | 1106 | (45) | 1.00 (0.88, 1.14) | 0.98 | -- | -- |
| >34 | 72 | (51) | 1056 | (43) | 0.98 (0.86, 1.13) | 0.82 | -- | -- |
| Income ≥10,000 KSH/month | 73 | (52) | 1051 | (43) | 0.98 (0.91, 1.06) | 0.65 | -- | -- |
| At least secondary education | 71 | (50) | 1129 | (46) | 0.99 (0.92, 1.07) | 0.86 | -- | -- |
| Married/cohabitating | 104 | (74) | 1608 | (66) | 0.98 (0.90, 1.07) | 0.68 | -- | -- |
| Contraception use ≥1 year | 51 | (37) | 1500 | (63) | 1.07 (0.98, 1.16) | 0.13 | -- | -- |

Table 3l: Concordance between preference for and use of non-hormonal methods

| Method attribute preferences and potential confounders | Use of hormonal method (n=1590) | | Use of non-hormonal method (n=988) | | Poisson generalized linear models | | | |
|---|---|---|---|---|---|---|---|---|
| | **N** | **(%)** | **N** | **(%)** | **Crude PR (95% CI)** | **p** | **Adjusted PR (95% CI)** | **p** |
| *Method attribute preference* | | | | | | | | |
| Non-hormonal | 777 | (49) | 556 | (56) | 1.20 (1.06, 1.36) | **0.004** | 1.20 (1.05, 1.36) | **0.007** |
| *Demographic characteristics* | | | | | | | | |
| Age | | | | | | | | |
| <25 | 177 | (11) | 110 | (11) | Ref. | Ref. | Ref. | Ref. |
| 25-34 | 792 | (50) | 372 | (38) | 0.83 (0.68, 1.04) | **0.09** | 0.84 (0.67, 1.05) | 0.12 |
| >34 | 621 | (39) | 506 | (51) | 1.18 (0.96, 1.45) | 0.12 | 1.13 (0.91, 1.41) | 0.29 |
| Income ≥10,000 KSH/month | 644 | (41) | 486 | (49) | 1.23 (1.09, 1.40) | **<0.001** | 1.21 (1.06, 1.37) | **0.004** |
| At least secondary education | 733 | (46) | 470 | (48) | 1.04 (0.92, 1.18) | 0.51 | -- | -- |
| Married/cohabitating | 1089 | (68) | 625 | (63) | 0.87 (0.76, 0.99) | **0.03** | 0.93 (0.81, 1.06) | 0.26 |
| Contraception use ≥1 year | 900 | (57) | 653 | (70) | 1.44 (1.25, 1.65) | **<0.001** | 1.36 (1.18, 1.56) | **<0.001** |
